# Global inequality in disability-adjusted life years due to eye diseases: a cross-national analysis from 1990 through 2015

**DOI:** 10.1101/2020.11.22.20236349

**Authors:** Wei Wang, Yingfeng Zheng, Wenyong Huang, Xiaoling Liang, Yizhi Liu

## Abstract

**Purpose:** The aim of this study was to evaluate the trends in global inequality in eye health by using data on the global burden of disease measured in disability-adjusted life years (DALYs).

**Methods:** This is an international observational study. We estimated the burden of eye disease by calculating the sum of DALYs (from the Global Burden of Disease study, 2016 update) due to cataract, refraction and accommodation disorders (RE), glaucoma, age-related macular degeneration (AMD), and other causes. We assessed the burden of various eye disease in relation to economic status and etiology by calculating the Gini coefficient, and the concentration index.

**Results:** This study included 195 countries, VI accounted for 2.46 (95%CI: 2.26 to 2.70) billion DALYs. Global inequality in the health burden due to VI has not changed greatly in the last 25 years, with a Gini coefficient of 0.217 in 1990 and 0.201 in 2015 Cataract was concentrated in poor socioeconomic countries, while AMD was concentrated in higher socioeconomic countries. In 1990, cataract was the most unevenly distributed cause, followed by AMD, RE, glaucoma, and other causes. In 2015, cataract remained the most unevenly distributed cause on a global scale, followed by RE, AMD, glaucoma, and other causes. Time trends of inequality in high-income economies differed from those in low- and middle-income economies. There is an ongoing reduction in the inequality of the VI burden across high-income countries; however, there is an upward trend across low- and middle-income countries. The inequality of RE has improved among high-, low-, and middle-income economies.

**Conclusion:** The health burden of eye diseases has not improved and that the global inequality of eye health has increased continuously in the last 25 years. Time trends of inequality in high-income economies differed from those in low- and middle-income economies.

## Introduction

Globally, 441 million people experienced some form of visual impairment (VI) in 2015, including 36.0 million people who were blind, 216.6 million moderate to severe visual impairment (MSVI).^1^ VI poses substantial economic burdens on societies and families.^2^ The total global cost of VI reached $3 trillion in 2010 and is estimated to increase by about 20% by 2020.^3^ Apart from visual difficulty, VI was also associated with loss of quality of life and increased risk for falls and mortality.^4, 5^ The disability-adjusted life year (DALY) is a single measure for evaluating disease burden across diseases and injuries, which is calculated by combining the number of healthy years lost due to premature death and the number of years living with disability.

Health inequality was defined as the unjust disparities in health among population due to poverty, lack of access to service, discrimination, unaffordability etc. Health inequality is an indicator for evaluating observable, measurable, and monitorable differences among sub-populations with the aim to reduce unfairness related to health. Gini coefficient and concentration index were the most commonly used metrics for quantifying health inequality. The global inequality of VI is a key consideration in the reduction of avoidable blindness, and identifying factors that contribute to health inequality is informative for policy planning in the context of limited resource allocation.^6, 7^ Inequality can be divided into intra-country inequalities and inter-country inequalities, with the former was more observed at home for individual. Even in developed countries the intra-country differences are often substantial. Using data from the Global Burden of Disease (GBD) 2004 study, Ono et al.^8^ found that cataract showed the widest variations from country to country. The latest GBD 2016 study evaluated the DALYs of 333 diseases and injuries across 195 countries/territories.^2^ However, the current status of inequality of VI was not analyzed by the GBD 2016 study.

Socioeconomic development differs across countries, which might play an important role in the global inequality of VI. Our previous studies demonstrated that socioeconomic factors explained over 70% of the variations in the prevalence of VI, cataract surgical rate (CSR) and cataract surgical coverage (CSC).^9-11^ The inequality of eye health may be a reflection of multi-dimensional poverty, including poor health, lack of education, etc. The past decades have witnessed socioeconomic progress in many countries, implementations of VISION 2020, and many other national programs.^12^ However, the time trends of global inequality remain unclear. We hypothesize that the global inequality of eye health has been improved in the last 25 years. The objective of this study was to use the latest GBD 2016 study to evaluate the trends over time of inequality in the health burden due to eye diseases.

## Methods

### Data sources

This is an international, time-series observational study. In 1993, the GBD 1990 study first introduced the DALY as a single measure for quantifying disease burden. The DALY is calculated based on the number of years of healthy life lost due to premature death and the number of years lived in inadequate health, a calculation that has been detailed in previous publications.^13, 14^ The latest GBD 2016 study has synthesized epidemiological data to produce DALY estimated by causes for 195 countries/territories around the world. The age-standardized DALY rates (per 100,000) for eye diseases in 1990, 1995, 2000, 2005, 2010, and 2015 were obtained from the GBD 2016 open database.^2^ The eye diseases were categorized into five subgroups: cataract, refraction and accommodation disorders (RE), glaucoma, age-related macular degeneration (AMD), and other causes. The Human Development Index (HDI) is a single measure of socioeconomic development, which reflects comprehensive attainment in health, education, and living standard in a region. The corresponding HDI in each country was obtained from the database of the United Nations Development Programme (UNDP) (http://hdr.undp.org/en/data).

### Health inequality indicators

Two health inequality indicators were adopted in this study: the Gini coefficient and the concentration index. The principle behind these indicators has been detailed in previous publications and summarized by the World Bank.^6-8, 15^ In brief, the Lorenz curve was constructed to calculate the Gini coefficient, which is the most common indicator of inequality. It assesses absolute inequality, which ranges from 0 (complete evenness) to 1 (complete unevenness). The Lorenz curve maps the cumulative fraction of the age-standardized DALY rate against the cumulative country proportion. The concentration index was obtained through the concentration curve, which shows the variability of the age-standardized DALY rate across countries ranked by HDI or gross domestic product (GDP) per capita. The concentration index is a relative measure of eye health inequality due to socioeconomic development, which reflects the extent to which the age-standardized DALY rate is concentrated among high- or low-income regions. A positive value indicates concentration among high-income countries, and a negative value indicates concentration among low-income countries. This study used the absolute value of the concentration index as ranging from 0 (no inequality due to socioeconomic development) to 1 (complete inequality due to socioeconomic development).

### Statistical analyses

Data analyses were performed by using Stata 12.0 SE (StataCorp LP, College Station, TX). Linear regression analysis was used to explore the impact of HDI on age-standardized DALY rate. The age-standardized DALY rate in each country was used for calculating Gini coefficient from 1990 to 2015. The age-standardized DALY rate and HDI in each country were used for calculating concentration index from 1990 to 2015. Linear regression analysis was used to evaluate the association between HDI and age-standardized DALY rate. A higher value of Gini coefficient and absolute value of concentration index indicate severer inequality. Line plots were constructed to show time trends of inequalities by using Microsoft Excel 2010 (Microsoft office, USA). A P less than 0.05 was considered as statistically significant.

## Results

This study included 195 countries, which are home to >95% of the world’s population. VI accounted for 2.46 (95%CI: 2.26 to 2.70) billion DALYs, with 14.60 (95%CI: 9.39 to 22. 94) million for RE, 3.88 (95%CI: 2.77 to 5.23) million for cataract, 541.3 (95%CI: 369.5 to 748.0) thousand for glaucoma, 462.4 (95%CI: 327.4 to 633.5) thousand for AMD, and 1.76 (95%CI: 1.25 to 2.39) million for other causes.

The age-standardized DALY rate due to RE decreased between 1990 and 2015 (Figure 1). The age-standardized DALY rate due to cataract showed an increasing trend between 1990 and 2005 and a stabilized trend between 2005 and 2015. From 1990 to 2015, the age-standardized DALY rate due to AMD and glaucoma showed a slight increase.

**Figure 1.**
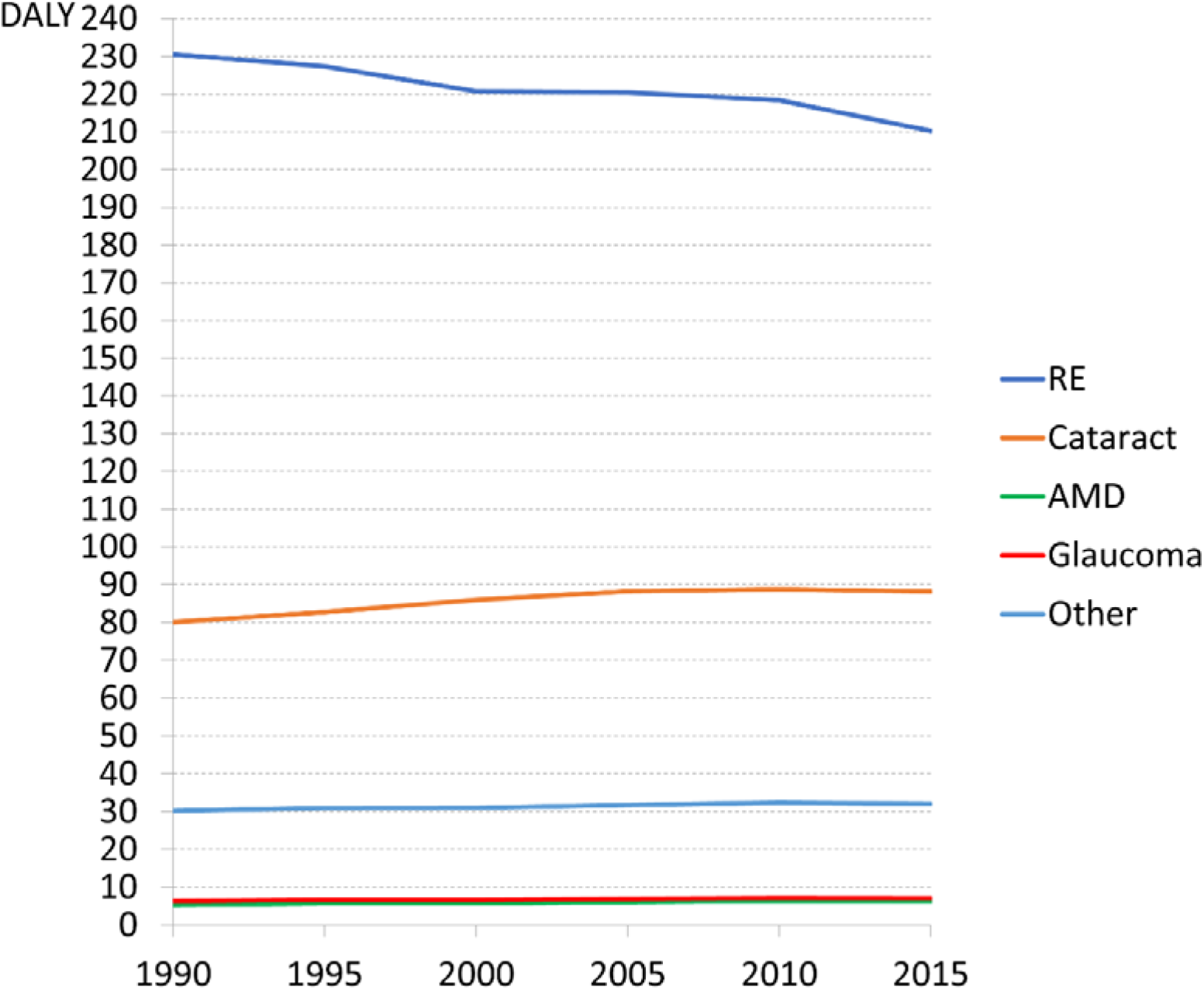
Trends over time in age-standardized DALY rates of vision loss and various causes worldwide. RE, vision loss caused by uncorrected refraction and accommodation disorders; cataract, vision loss caused by cataract; glaucoma, vision loss caused by glaucoma; AMD, vision loss caused by age-related macular degeneration; other causes, vision loss due to other causes.

Figure 2 shows the variability of DALY rate of VI in 2015 across countries. Analyses of associations between the age-standardized DALY rate of VI and HDI showed negative correlations (Figure 3). With the growth in HDI, the age-standardized DALY rate decreased (all P<0.001).

**Figure 2.**
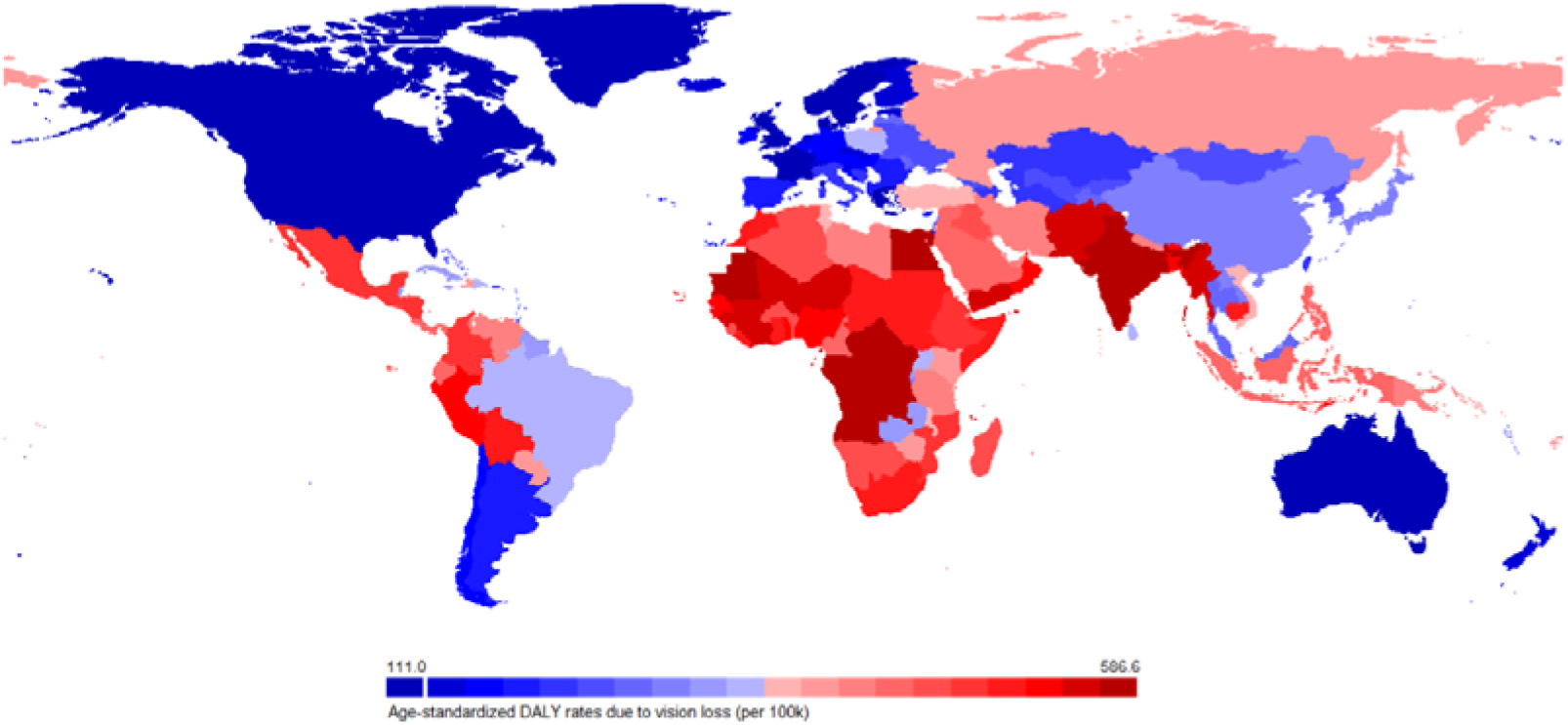
World-map of DALY rate due to overall vision loss across countries in 2015.

**Figure 3.**
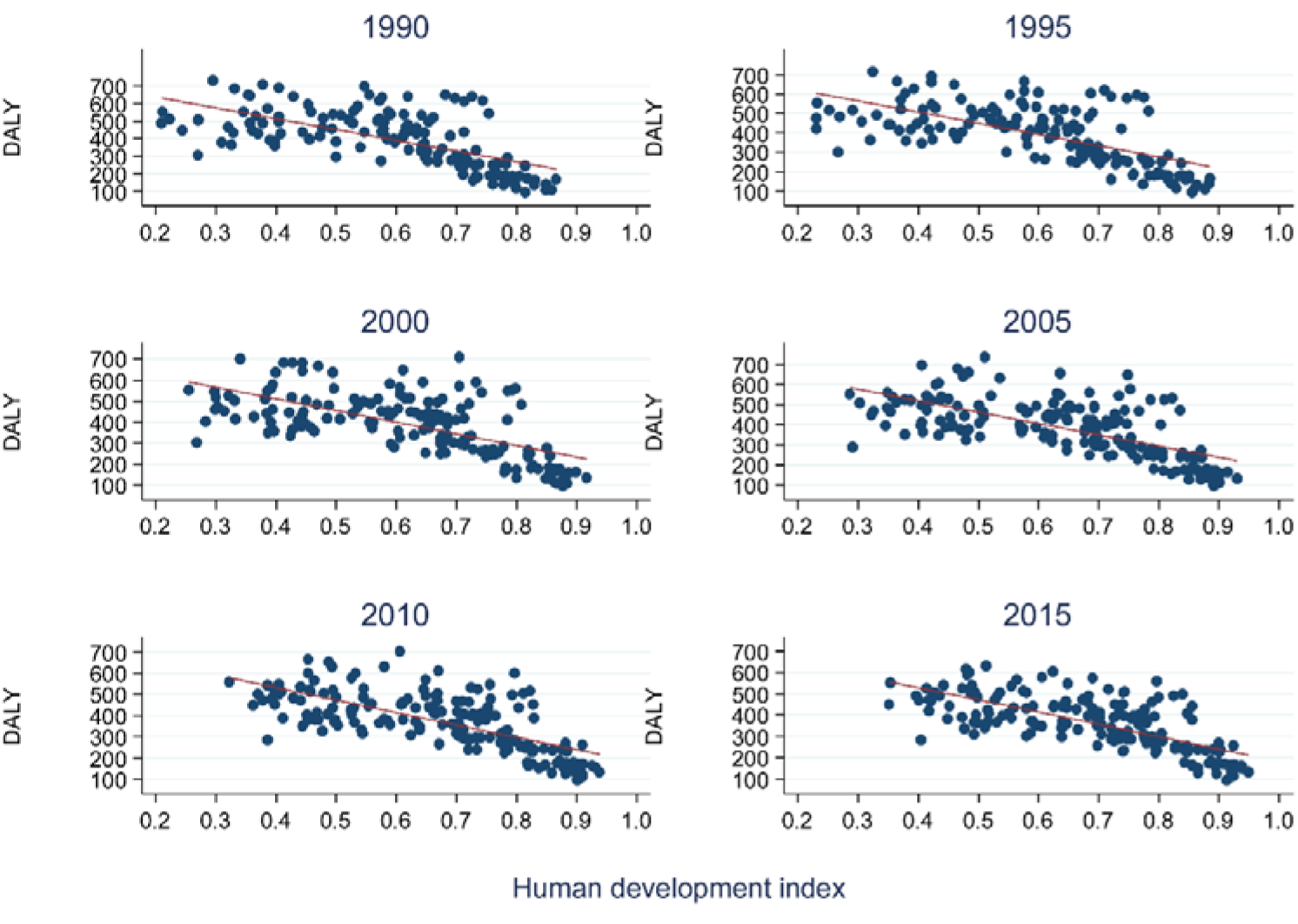
The relationship between age-standardized DALY rates and human development index from 1990 to 2015.

Global inequality in the health burden due to VI has not changed greatly in the last 25 years, with a Gini coefficient of 0.217 in 1990 and 0.201 in 2015 (Figure 4A). The Gini coefficient of cataract decreased between 1990 and 1995 and then increased to an equivalent level in 2000. Both the Gini coefficient of the glaucoma and RE decreased consistently from 1990 to 2015. An analysis of the concentration index of VI demonstrated an increasing trend between 1990 and 2005, with the level being maintained between 2005 and 2015. From 1990 to 2015, the concentration index increased continuously for cataract, glaucoma, and AMD, and reduced for RE. Cataract was concentrated in poor socioeconomic countries, while AMD was concentrated in higher socioeconomic countries. In 1990, cataract was the most unevenly distributed cause, followed by RE, AMD, glaucoma, and other causes. In 2015, cataract remained the most unevenly distributed cause on a global scale, followed by other causes, RE, AMD, glaucoma (Figure 4B).

**Figure 4.**
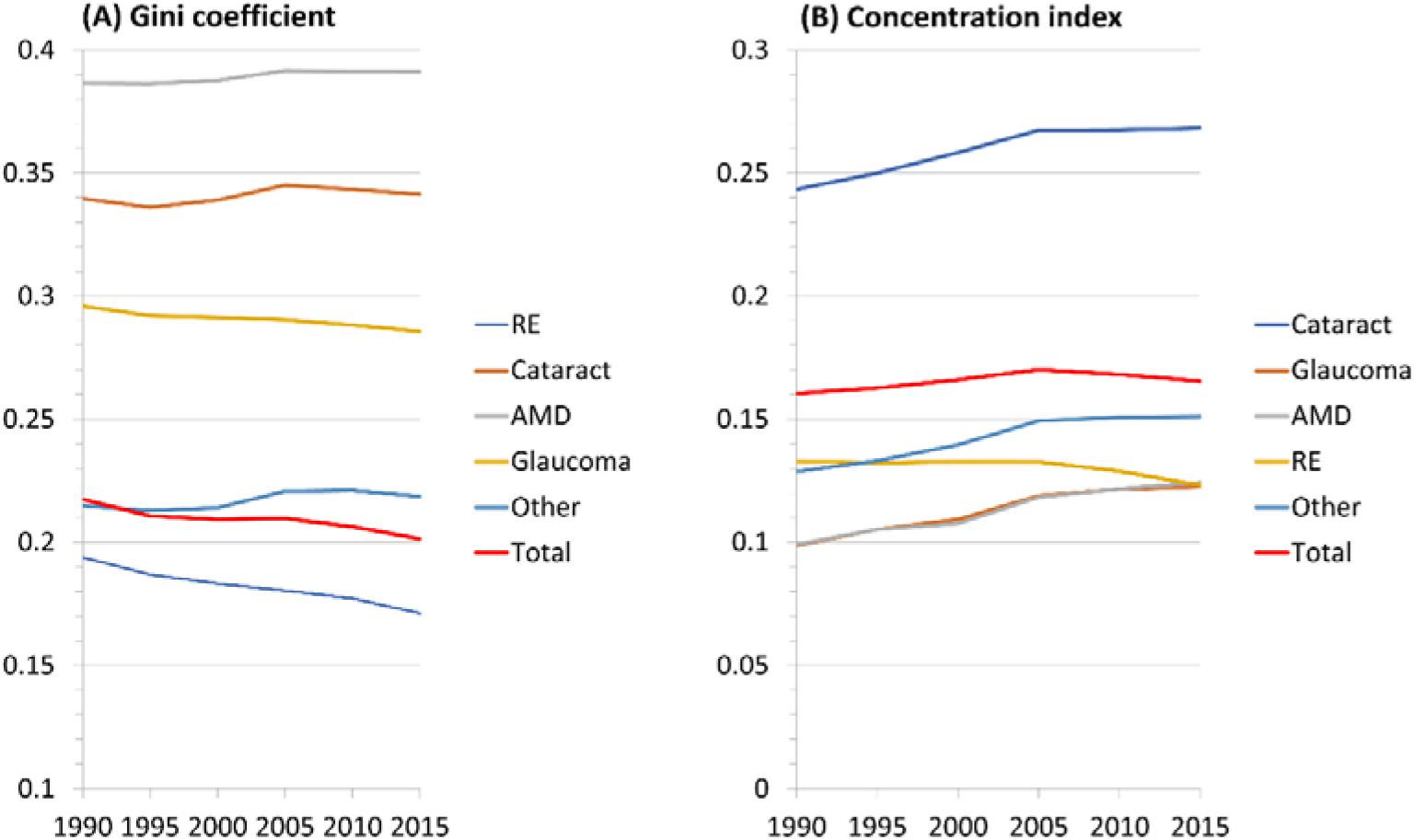
Trends of Gini coefficients and concentration index by specific causes from 1990 to 2015.

Figure 4 shows the Gini coefficient of age-standardized DALY rates by specific cause and economic level. The inequality of all diseases has been improved across high-income countries. Across high-income countries, cataract was most unevenly distributed, followed by glaucoma, AMD, and RE (Figure 5A). Across low- and middle-income countries, the overall inequality of VI did not improve (Figure 5B). The inequality of RE improved dramatically, from the second-ranking unevenly distributed disease in 1990 to the third-ranking in 2015. The inequality of other diseases did not change obviously over the past 25 years. Sensitivity analysis based on 95% prediction interval was presented in supplementary table 1. The intra-country inequalities of age-standardized DALY rate due to ocular diseases in USA was presented in supplementary table 2.

**Figure 5.**
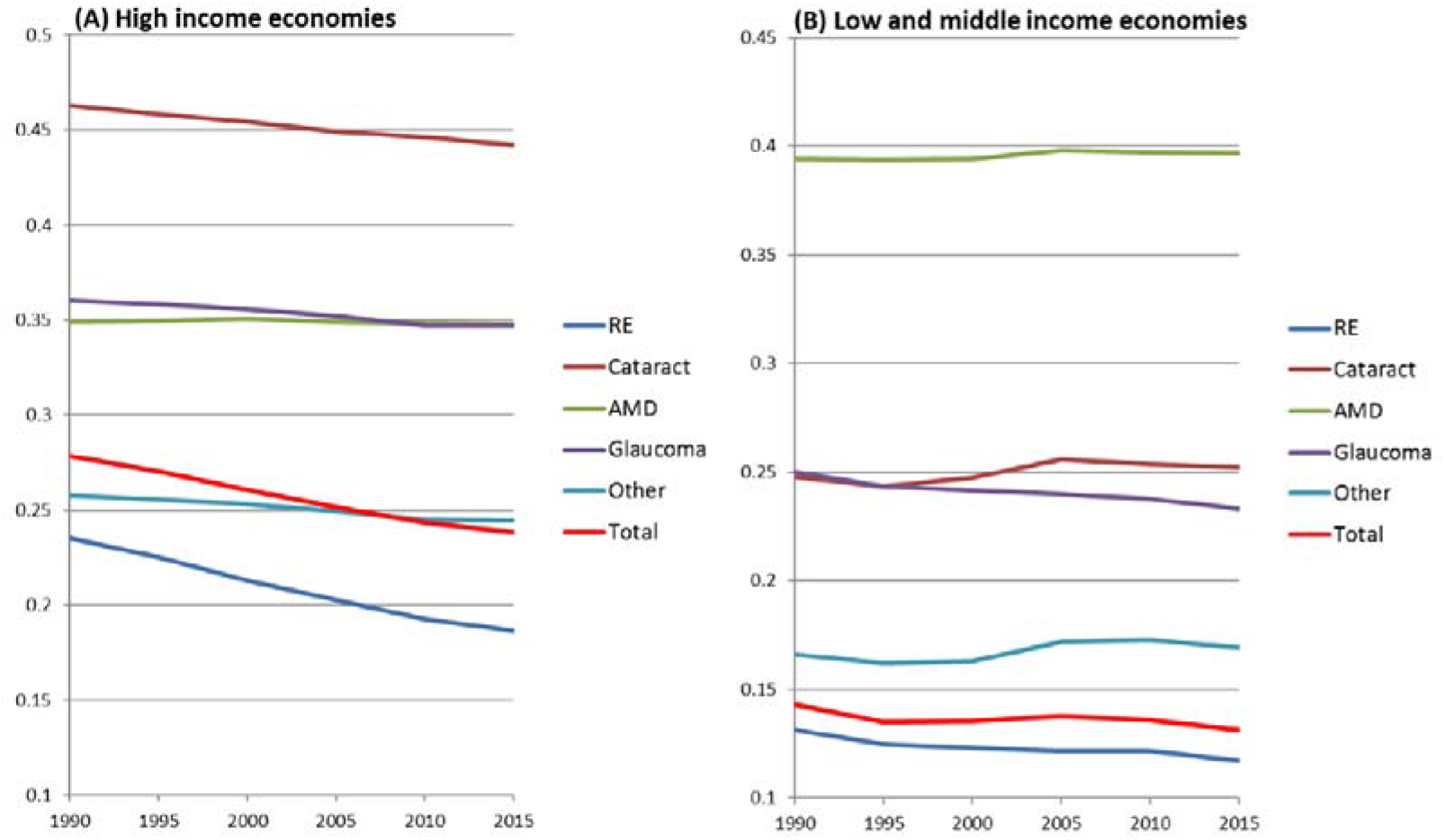
Trends of Gini coefficients by specific causes of vision loss and socioeconomic status from 1990 to 2015.

Trend analyses of the concentration index revealed a reduction in socioeconomic disparities in overall VI across high-income countries (Figure 6A). Among the specific causes, the inequality of cataract, AMD, glaucoma and RE decreased, and the inequality of AMD did not change dramatically. Across low- and middle-income countries, overall socioeconomic disparities have increased since 1990, reaching a maximum in 2005 (Figure 6B). Among specific causes, the inequality of cataract and AMD increased, and the inequality of RE and glaucoma did not change dramatically.

**Figure 6.**
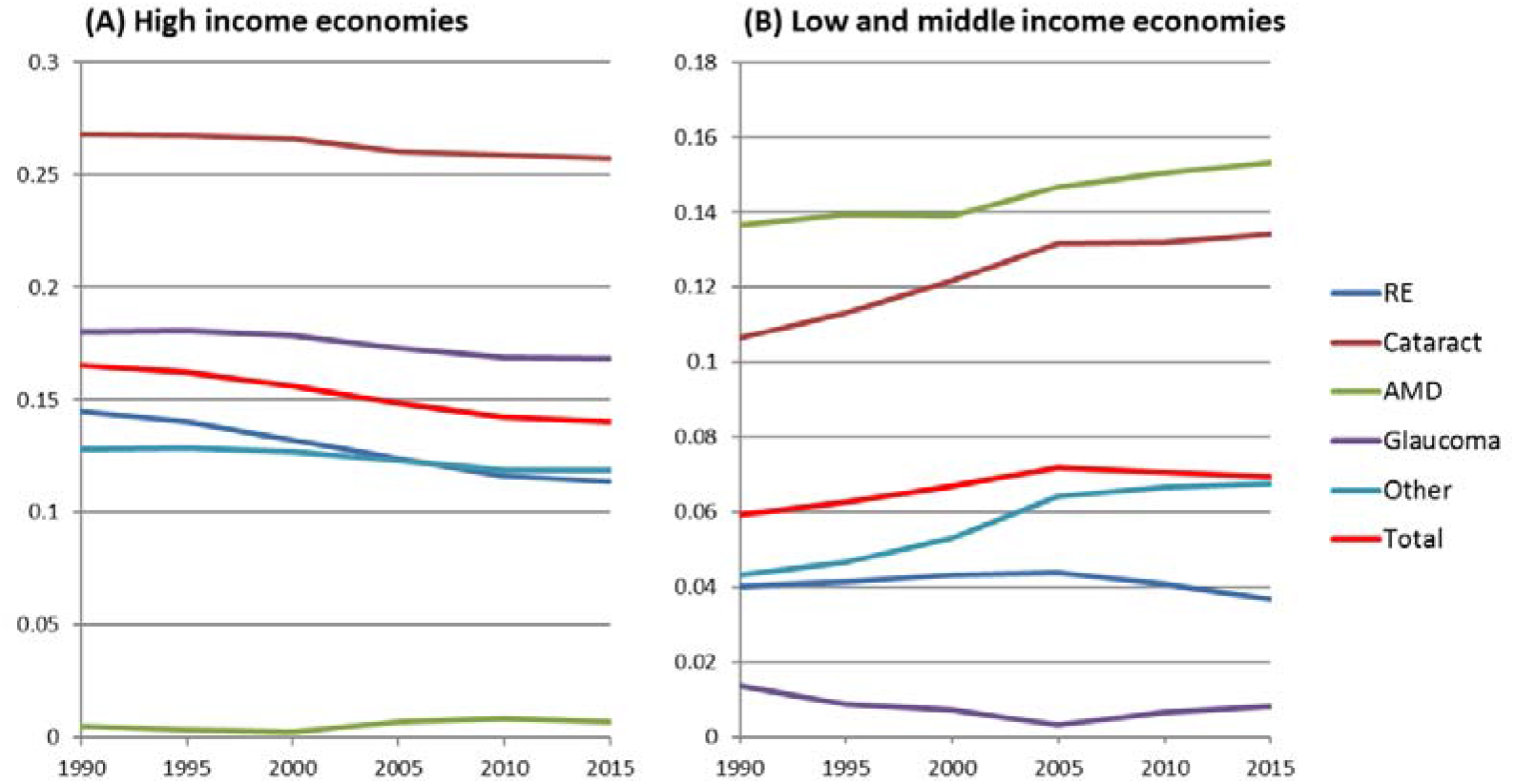
Trends of concentration index by specific causes of vision loss and socioeconomic status from 1990 to 2015.

## Discussion

This study demonstrated that the overall inequality in eye health has not improved on the global scale in the last 25 years. Time trends of inequality in high-income economies differed from those in low- and middle-income economies. There is an ongoing reduction in the inequality of the VI burden across high-income countries; however, there is an upward trend across low- and middle-income countries, particularly for cataract and AMD.

International statistics on VI may be presented from different points of view. The “eye health” perspective represents the medical point of view, and compares different causes of vision loss. The “disability” perspective (DALYs) represents the point of view of individuals and their quality of life. DALYs are based on a presumed trade-off between quality and quantity of life (How many years would you trade for perfect health?). Finally, there is the “socio-economic” point of view, which measures the cost to society. It is important to realize that these points of view are related, but that they are not interchangeable. In this study, we estimated the time trends of inequality of DALY rate between 1990 and 2015 in the perspective of country-to-country differences. This additional point of view is helpful for understanding the current statues of preventing blindness across countries. Inter-country variability may increase when some countries progress more than others, but also when most progress, but some stagnate. The inequality scores alone, cannot distinguish between these cases. The outliers are informative for policy planning. For example, the countries with greater progress can be serve as samples, while the countries with lowest progress warranted more invest and support from international society. Future study is needed to report on the outliers, when adequate data are available.

The age-standardized DALY rates of all eye diseases did not reduce between 1990 and 2015. The latest “Universal Eye Health: The Global Action 2014–2019” set a new target: to reduce the prevalence of VI before the end of 2019 by 25% compared to the 2010 baseline.^16^ Past decades witnessed the improvement in blindness prevalence and lack of an absolute reduction of blindness people, which may be related to population increases and increases in longevity. From 1990 to 2010, the age-standardized prevalence of older adults with blindness decreased from 3.0% in 1990 to 1.9% in 2010.^17, 18^ However, the number of people with blindness increased by 17.6% from 1990 to 2015, which was attributed to population growth (38.4%) and population aging (34.6%) worldwide.^1^ This study further revealed that the age-standardized DALY rates of all eye diseases did not change, which may be associated with population growth and extended life expectancy. The average life expectancy increased from 66.4 in 1990 to 71.4 in 2015, leading to greater loss of a healthy life at the same onset age.^19^

By using the crude DALY rates (DALYs per 100,000 population) in 2004, Ono et al.^8^ reported that the most unevenly distributed cause was cataract, followed by the RE, glaucoma, and AMD, on the global scale. Across high-income countries, cataract was distributed most unevenly, followed by RE, glaucoma, and AMD. Across low- and middle-income countries, RE was distributed most unevenly, followed by cataract, glaucoma, and AMD. We adopted the age-standardized DALY rate in this study instead of the crude DALY rate because it adjusted the age structure and was a more appropriate measure for inter-country comparisons. By using data from the GBD 2013 study, Lou et al.^15^ (2017) recently reported that the socioeconomic inequality in the cataract burden increased between 1990 and 2013. We confirmed the finding and conducted a further investigation to assess health inequality in specific eye diseases from 1990 to 2015 by socioeconomic development. Apart from cataract, health inequality exists in other eye diseases. Inequality across high-income countries has been improving, while that across low- and middle-income countries has been rising. Compared to earlier GBD studies, the estimates presented in the GBD 2016 study were more accurate because of the inclusion of the near VI estimation.

Eye health was closely associated with socioeconomic development. It was reported that the utility of eye care was significantly associated with education, race, and socioeconomic status in the US, with a higher proportion of individuals with less education and lower income being less able to afford eyeglasses.^20^ Limited studies were available on the impact of socioeconomic development on eye health on a global scale. It was estimated that the prevalence of blindness reduced dramatically when the national income per capita increased from USD$10,000 to USD$20,000 in a country.^21^ Evidence showed that HDI and GNI per capita were significantly associated with the cutoff of visual acuity for cataract surgery.^22^ An analysis of data from 38 countries showed that the annual rate of glaucoma surgery per million people was associated with GDP and the number of ophthalmologists.^23^ Our previous studies demonstrated that the HDI and GDP per capita were independently associated with the quantity and quality of cataract surgery in a country.^9, 10^

We observed that the rate of progress in different eye disease were different, which may be related to the very different approaches for different causes of VI. Cataract requires surgical facilities and trained surgeons. Glaucoma requires early detection, which might be performed by technicians. RE requires screening, particularly of children, which can often be performed by volunteers. It also requires the general availability of glasses, which often is a problem of infrastructure, rather than of medical care. AMD is not treatable, except with expensive medications, which implies reliance on economic resources as well as on trained clinicians. That there are relatively few remarkable differences, may point to the fact that they are all dependent on socio-economic development.

Cataract ranks as the most unevenly distributed eye disease. The health inequality of cataract increased consistently from 1990 to 2015 in both developed and developing countries. There may be different reasons explaining the rise in developed and developing countries. In developed countries, more people with good preoperative visual acuity received cataract surgery.^24, 25^ The cataract surgical rates are steadily increased, which may be partly explained by an aging demographic structure, reduced VI thresholds as an indication for surgery, increased frequency of second eye surgery, and increasing expectations by patients for better vision. ^24, 25^ However, not all high-income countries are equipped with full insurance coverage, adequate health expenditure, and perfect basic health services. In developing countries, vast needs for cataract surgery have not been fulfilled.

It is noteworthy that the health inequality of RE has continued to reduce in the last 25 years. The interest in uncorrected RE is relatively recent. The interest in presbyopia is even more recent. Both developed countries and developing countries showed reducing trends. The changes for RE may be in part due to better surveillance. The socioeconomic development may also contribute to the changes, as shown in Figure 5, the inequality of RE in developed countries decreased greater than that in developing countries. These observations, including a very large proportion of uncorrected presbyopia cases, highlight the need to scale up VI alleviation efforts at all levels.^26^

This study does have some limitations. First, there are limitations in the GBD 2016 study, such as statistical assumptions, inclusion of self-reported data, the lack of epidemiological surveys in some parts of the world. However, the GBD 2016 study provides the most updated and accurate estimates of DALY rates across 195 countries. This study examines inequalities in VI among countries based on the estimates generated by the GBD model, assuming that these estimates are accurate, which is highly questionable. In addition, socioeconomic development indicators are used as covariates by the GBD mathematical model to predict estimates for countries and data points where there is no data. This creates a circular reasoning in the approach used by the authors. The association between prevalence of VI and socioeconomic development is inherent to the model. Second, the usage of data at the national level may introduce bias because variations exist across provinces in a country. However, summary data at provincial or district levels are usually not suited for international comparisons across countries. Third, other major vision-threatening diseases, such as diabetic retinopathy, were not analyzed as a separate category and were combined in the other causes. Further studies based on individual level data are needed to explore the efficacy of policy interventions on socioeconomic inequality in the health burden of vision loss. Finally, inequalities also exist within countries. These inequalities are much more important from a public health perspective because they can be addressed through appropriate policies. Inequalities among countries cannot be easily addressed because there are no global governance mechanisms that could allocate more resources to eye care. However, only USA data across states were available for inequality estimation. International platform collecting provincial data across countries are helpful to guide the future global actions.

In summary, this study found that the health burden of eye diseases has not improved and that the global inequality of eye health has increased continuously in the last 25 years. In general, health inequality related to socioeconomic development showed an ongoing increasing trend for all categories of VI, particularly for cataract and glaucoma. The global inequality of RE did not change substantially, while that of AMD decreased. Time trends of inequality in high-income economies differed from those in low- and middle-income economies. These findings highlight the urgent need for the provision of more eye care services and infrastructure in developing countries to achieve GAP’s target before 2020.

## Data Availability

The availability of all data referred to in the manuscript and note links below.

## Acknowledgments

Support was provided by the Guangdong Province Science & Technology Plan (2014B020228002).

## References

1. Bourne R, Flaxman SR, Braithwaite T, Cicinelli MV, Das A, Jonas JB, Keeffe J, Kempen JH, Leasher J, Limburg H, et al. Magnitude, temporal trends, and projections of the global prevalence of blindness and distance and near vision impairment: A systematic review and meta-analysis. Lancet Glob Health 2017;5:e888–97.

2. GBD 2016 DALYs and HALE Collaborators. Global, regional, and national disability-adjusted life-years (DALYs) for 333 diseases and injuries and healthy life expectancy (HALE) for 195 countries and territories, 1990-2016: A systematic analysis for the Global Burden of Disease Study 2016. Lancet 2017;390:1260–344.

3. Gordois A, Cutler H, Pezzullo L, Gordon K, Cruess A, Winyard S, Hamilton W, Chua K. An estimation of the worldwide economic and health burden of visual impairment. Glob Public Health 2012;7:465–81.

4. Cugati S, Wang JJ, Knudtson MD, Rochtchina E, Klein R, Klein BE, Wong TY, Mitchell P. Retinal vein occlusion and vascular mortality: Pooled data analysis of 2 population-based cohorts. Ophthalmology 2007;114:520–4.

5. Xu L, Wang YX, Wang J, Jonas JJ. Mortality and ocular diseases: The Beijing Eye Study. Ophthalmology 2009;116:732–8.

6. Barr B, Higgerson J, Whitehead M. Investigating the impact of the English health inequalities strategy: Time trend analysis. BMJ 2017;358:j3310.

7. Barr B, Bambra C, Whitehead M. The impact of NHS resource allocation policy on health inequalities in England 2001-11: Longitudinal ecological study. BMJ 2014;348:g3231.

8. Ono K, Hiratsuka Y, Murakami A. Global inequality in eye health: Country-level analysis from the Global Burden of Disease Study. Am J Public Health 2010;100:1784–8.

9. Wang W, Yan W, Muller A, He M. A global view on output and outcomes of cataract surgery with national indices of socioeconomic development. Invest Ophthalmol Vis Sci 2017;58:3669–76.

10. Wang W, Yan W, Fotis K, Prasad NM, Lansingh VC, Taylor HR, Finger RP, Facciolo D, He M. Cataract surgical rate and socioeconomics: A global study. Invest Ophthalmol Vis Sci 2016;57:5872–81.

11. Wang W, Yan W, Müller A, Keel SE, He M. Socioeconomics and prevalence of visual impairment and blindness: A global analysis. Jama Ophthalmol 2017;135:1–8.

12. Wong TY, Zheng Y, Jonas JB, Flaxman SR, Keeffe J, Leasher J, Naidoo K, Pesudovs K, Price H, White RA, et al. Prevalence and causes of vision loss in East Asia: 1990-2010. Br J Ophthalmol 2014;98:599–604.

13. GBD 2015 DALYs and HALE Collaborators. Global, regional, and national disability-adjusted life-years (DALYs) for 315 diseases and injuries and healthy life expectancy (HALE), 1990-2015: A systematic analysis for the Global Burden of Disease Study 2015. Lancet 2016;388:1603–58.

14. GBD 2015 Disease and Injury Incidence and Prevalence Collaborators. Global, regional, and national incidence, prevalence, and years lived with disability for 310 diseases and injuries, 1990-2015: A systematic analysis for the Global Burden of Disease Study 2015. Lancet 2016;388:1545–602.

15. Lou L, Wang J, Xu P, Ye X, Ye J. Socioeconomic Disparity in Global Burden of Cataract: An Analysis for 2013 with Time Trends Since 1990. Am J Ophthalmol 2017;180:91–6.

16. World Health Organization (WHO). Universal eye health: A global action plan 2014-2019. Available from http://www.who.int/blindness/actionplan/en/. Accessed April 1, 2019. 2013.

17. Yan X, Guan C, Mueller A, Iezzi B, He M, Liang H, Meltzer M, Congdon NG. Outcomes and projected impact on vision restoration of the China Million Cataract Surgeries Program. Ophthalmic Epidemiol 2013;20:294–300.

18. Stevens GA, White RA, Flaxman SR, Price H, Jonas JB, Keeffe J, Leasher J, Naidoo K, Pesudovs K, Resnikoff S, et al. Global prevalence of vision impairment and blindness: Magnitude and temporal trends, 1990-2010. Ophthalmology 2013;120:2377–84.

19. He M, Wang W, Huang W. Variations and trends in health burden of visual impairment due to cataract: A global analysis. Invest Ophthalmol Vis Sci 2017;58:4299–306.

20. Zhang X, Cotch MF, Ryskulova A, Primo SA, Nair P, Chou CF, Geiss LS, Barker LE, Elliott AF, Crews JE, et al. Vision health disparities in the United States by race/ethnicity, education, and economic status: Findings from two nationally representative surveys. Am J Ophthalmol 2012;154:S53–62.

21. Ho VH, Schwab IR. Social economic development in the prevention of global blindness. Br J Ophthalmol 2001;85:653–7.

22. Lansingh VC, Carter MJ. Use of Global Visual Acuity Data in a time trade-off approach to calculate the cost utility of cataract surgery. Arch Ophthalmol 2009;127:1183–93.

23. Mansouri K, Medeiros FA, Weinreb RN. Global rates of glaucoma surgery. Graefes Arch Clin Exp Ophthalmol 2013;251:2609–15.

24. Lundstrom M, Goh PP, Henry Y, Salowi MA, Barry P, Manning S, Rosen P, Stenevi U. The changing pattern of cataract surgery indications: A 5-year study of 2 cataract surgery databases. Ophthalmology 2015;122:31–8.

25. Erie JC. Rising cataract surgery rates: Demand and supply. Ophthalmology 2014;121:2–4.

26. Frick KD, Joy SM, Wilson DA, Naidoo KS, Holden BA. The global burden of potential productivity loss from uncorrected presbyopia. Ophthalmology 2015;122:1706–10.

