## Supplemental Tables for "Global inequality in disability-adjusted life years due to eye diseases: a cross-national analysis from 1990 through 2015"

**Supplementary materials**

**sTable 1.** Sensitivity analysis of Gini coefficient for age-standardized DALY rates worldwide.

| **Gini coefficient** | **Worldwide** | | | | | |
|---|---|---|---|---|---|---|
| | **1990** | **1995** | **2000** | **2005** | **2010** | **2015** |
| **Upper estimation of age-standardized DALY rate** | | | | | | |
| RE | 0.195 | 0.189 | 0.184 | 0.182 | 0.179 | 0.173 |
| Cataract | 0.338 | 0.335 | 0.338 | 0.343 | 0.342 | 0.343 |
| AMD | 0.386 | 0.386 | 0.386 | 0.391 | 0.390 | 0.391 |
| Glaucoma | 0.294 | 0.289 | 0.289 | 0.288 | 0.286 | 0.286 |
| Other | 0.214 | 0.212 | 0.214 | 0.221 | 0.221 | 0.218 |
| Total | 0.214 | 0.207 | 0.205 | 0.205 | 0.202 | 0.199 |
| **Lower estimation of age-standardized DALY rate** | | | | | | |
| RE | 0.196 | 0.190 | 0.186 | 0.183 | 0.179 | 0.173 |
| Cataract | 0.342 | 0.338 | 0.342 | 0.348 | 0.345 | 0.338 |
| AMD | 0.388 | 0.388 | 0.389 | 0.394 | 0.394 | 0.393 |
| Glaucoma | 0.299 | 0.294 | 0.294 | 0.293 | 0.291 | 0.289 |
| Other | 0.217 | 0.215 | 0.215 | 0.222 | 0.223 | 0.214 |
| Total | 0.224 | 0.218 | 0.216 | 0.217 | 0.213 | 0.207 |

DALY, disability-adjusted life year; RE, refraction and accommodation disorders; AMD, age-related macular degeneration (AMD).

**sTable 2.** The intra-country inequalities of age-standardized DALY rate due to ocular diseases in USA.

| Gini coefficient | USA | | | | | |
|---|---|---|---|---|---|---|
| | **1990** | **1995** | **2000** | **2005** | **2010** | **2015** |
| RE | 0.011 | 0.011 | 0.011 | 0.011 | 0.010 | 0.010 |
| Cataract | 0.012 | 0.013 | 0.013 | 0.014 | 0.017 | 0.016 |
| AMD | 0.011 | 0.011 | 0.012 | 0.013 | 0.016 | 0.015 |
| Glaucoma | 0.011 | 0.012 | 0.012 | 0.013 | 0.017 | 0.016 |
| Other | 0.017 | 0.018 | 0.018 | 0.019 | 0.021 | 0.019 |
| Total | 0.008 | 0.009 | 0.009 | 0.009 | 0.009 | 0.008 |

DALY, disability-adjusted life year; RE, refraction and accommodation disorders; AMD, age-related macular degeneration (AMD).
